# Interrelations of aortic spring function, cardiovascular disease risk factors, and left ventricular diastolic function: The Framingham Heart Study

**DOI:** 10.1101/2025.06.30.25330569

**Authors:** Leroy L. Cooper, Brenton R. Prescott, Vanessa Xanthakis, Jian Rong, Martin G. Larson, Emelia J. Benjamin, Naomi M. Hamburg, Ramachandran S. Vasan, Gary F. Mitchell

**Author notes:** **Address for correspondence:** Leroy L. Cooper, PhD, MPH, Biology Department, Vassar College, 124 Raymond Ave., Box 70, Poughkeepsie, NY 12604.

## Abstract

**Background:** Energy associated with proximal aortic stretch during systole is recovered as diastolic elastic recoil of the aorta that facilitates left ventricular filling. Impairment of this aortic spring mechanism may contribute to left ventricular diastolic dysfunction. However, cross-sectional and longitudinal inter-relations of cardiovascular disease risk factors, aortic stretch, and left ventricular diastolic function have not been examined. The goal of this study was to assess the cross-sectional and longitudinal associations of cardiovascular disease risk factors and systolic atrioventricular plane displacement (AVPD), a surrogate measure of stretch of the mechanically coupled ascending aorta, with measures of left ventricular diastolic function.

**Methods:** At two examinations (14±1 years apart) in Framingham Heart Study participants (N=7117; mean age 50 years, 55% women), we assessed AVPD and left ventricular diastolic function using echocardiography. We measured systolic AVPD using the integral of the tissue Doppler s’ wave. Additionally, we assessed e’ (the peak early diastolic tissue velocity of the lateral mitral annulus) and E/e’ (the ratio of peak early mitral inflow velocity and e’).

**Results:** In cross-sectional analyses, higher AVPD was associated with higher e’ (*β* per SD±standard error=0.43±0.01; *P*<0.001) and lower E/e’ (*β*=-0.35±0.01; *P*<0.001). In longitudinal models (between two examinations), greater change in (Δ) AVPD between visits was associated with higher Δe’ (*β*=0.40±0.01; *P*<0.001) and lower ΔE/e’ (*β*=-0.23±0.01; *P*<0.001). We observed significant effect modification (interaction *P*-values: <0.001 to 0.045) for cross-sectional and longitudinal associations by median age, sex, obesity status, hypertension treatment, and the extent of aortic stiffness (assessed via carotid-femoral pulse wave velocity).

**Conclusion:** The aortic spring function, as assessed via AVPD, may play an important role in maintaining left ventricular diastolic function, with putative effects modified by aortic stiffness, obesity, age, and sex.

## Introduction

Diastolic dysfunction of the left ventricle (LV) is a common pathophysiologic condition that is characterized by impaired ventricular relaxation as well as elevated cardiac stiffness and ventricular filling pressures.^1^ It is widely believed that the mechanical twisting of the LV during systole and untwisting during diastole expels blood during contraction and generates suction during relaxation, aiding in rapid left ventricular filling.^2^ Additionally, abnormal hemodynamic coupling of the LV and aorta has long been recognized as a contributor to left ventricular diastolic dysfunction.^3^ However, research on the role of direct mechanical coupling of the LV and aorta (physical connection) and its role in left ventricular diastolic dysfunction and subsequent disease risk is limited.

In a sample of Framingham Heart Study (FHS) participants, we reported that aortic stiffening contributes a substantial and often overlooked mechanical load on the heart that is associated with lower left ventricular long-axis shortening.^4^ Importantly, we also observed that energy associated with stretch of the ascending aorta during systole can be recovered as elastic recoil during diastole (akin to a spring).^4^ During systole, the LV shortens longitudinally, displacing the atrioventricular plane towards the apex, which remains relatively stationary.^5–8^

The distal ascending aorta is relatively immobile during the cardiac cycle at the level of the brachiocephalic origin and beyond, whereas the aortic root and proximal ascending aorta are anchored to the highly mobile base of the heart. Systolic displacement of the atrioventricular plane toward the LV apex pulls on the aortic root and contributes to stretch of the ascending aorta during systole.^6,8^ Therefore, atrioventricular plane displacement (AVPD) not only indirectly indicates left ventricular systolic function but also captures the mechanical stretch of the proximal aorta associated with aortic spring function. Furthermore, we have posited that elastic recoil of the ascending aorta stretches the left ventricular long axis, which may contribute to the early diastolic suction that facilitates the transfer of blood from the left atrium into the LV.^8^

In a sample of Age, Gene/Environment Susceptibility (AGES)-Reykjavik participants, impairment of the aortic spring mechanism was associated with lower early left ventricular diastolic filling, particularly in women,^9^ which may contribute to higher risk for heart failure with preserved ejection fraction (HFpEF) in older women. In addition to these sex differences, aging and obesity also contribute to HFpEF risk and are associated with stiffening of the aorta.^10–12^ Although higher aortic stiffness may not be independently related to incident HFpEF,^13,14^ aortic stiffness may exacerbate the effects of existing risk factors and comorbidities that modify HFpEF risk.^15^ The foregoing AGES cross-sectional analyses raise the possibility that aortic stiffening could impair mechanical coupling of the LV and proximal aorta and contribute to left ventricular systolic and diastolic dysfunction. Therefore, analysis of potential effects of comorbid conditions on putative mechanisms contributing to left ventricular dysfunction and HFpEF could deepen our understanding of HFpEF susceptibility in individuals with prevalent risk factors.

The associations of longitudinal changes in cardiovascular disease (CVD) risk factors and measures of aortic spring function with longitudinal progression of left ventricular diastolic dysfunction have not been examined. Therefore, we investigated the relations of baseline and change in aortic spring function and left ventricular diastolic measures with baseline and change in CVD risk factors over a 14-year interval. We hypothesized that aortic stiffening increases the systolic load on the LV, particularly along its long axis, which reduces aortic longitudinal stretch during systole, reduces the potential energy stored in the aortic spring, and thereby impairs left ventricular diastolic function. To test this hypothesis, we assessed the cross-sectional and longitudinal associations of AVPD, a surrogate measure of aortic stretch, with measures of left ventricular diastolic function in a sample of FHS participants across the full adult age spectrum.

## Methods

Our study followed the Strengthening the Reporting of Observational Studies in Epidemiology (STROBE) reporting guidelines.^16^ The procedure for requesting data from the FHS can be found at https://www.framinghamheartstudy.org/.

### Study sample

The sample was drawn from the Framingham Offspring, New Offspring Spouse, Third Generation, and Omni-1 and Omni-2 cohorts, which have been described previously.^17–19^ Omni participants were Black, Asian, or Hispanic individuals who resided in the MetroWest area of Massachusetts. We included participants at two examination cycle visits at which arterial tonometry and echocardiography were routinely performed. At baseline (visit 1), Framingham Offspring (N=3021) and Omni-1 (N=298) participants at examination cycles 8 and 3, respectively, as well as Third Generation (N=4095), New Offspring Spouse (N=103), and Omni-2 (N=410) participants at examination cycle 1, were candidates for this investigation. However, 107 participants were ineligible due to offsite visits with limited examination. Of the 7820 eligible participants, we excluded participants who were missing data for independent or dependent variables (N=161) and covariate data (N=542) for the primary analyses. At visit 2, Framingham Offspring (N=1501) and Omni-1 (N=197) participants at examination 10 as well as Third Generation (N=3171), New Offspring Spouse (N=56), and Omni-2 (N=294) participants at examination 3 were candidates for this investigation. Due to coronavirus disease restrictions, 317 participants were ineligible due to remote or limited examinations. Of the 4902 eligible participants, we excluded participants who were missing data for independent or dependent variables (N=112) and covariate data (N=481) for the primary analyses. For the longitudinal analysis, we excluded the 258 participants who were not included in both visit 1 and 2 samples. All protocols were approved by Boston University Medical Center’s Institutional Review Board, and all participants provided written informed consent.

### Noninvasive echocardiographic and arterial tonometry

We performed a comprehensive hemodynamic assessment of the heart and aorta using arterial tonometry, echocardiography, and electrocardiography. We obtained arterial tonometry with simultaneous electrocardiography of the brachial, radial, femoral, and carotid arteries in supine participants using a custom tonometer as previously described.^20^ Next, all participants underwent echocardiographic evaluations, which included M-mode, 2-dimensional, pulsed-wave Doppler, and tissue Doppler imaging (Philips Healthcare, Andover, MA).^21^ We also obtained 2-dimensional echocardiographic images of the left ventricular outflow tract from a parasternal long-axis view followed by pulsed Doppler of the left ventricular outflow tract from an apical 5-chamber view. We digitized and transferred echocardiographic, tonometric, and electrocardiographic data to a core laboratory (Cardiovascular Engineering, Inc., Needham, MA) for blinded analyses.

### Assessment of left ventricular diastolic function and cardiac structure

We assessed mitral inflow and mitral annulus tissue Doppler in an apical 4-chamber view with participants in the left lateral decubitus position. We measured the peak early diastolic tissue velocity of the lateral mitral annulus (e’) and transmitral Doppler flow velocities (E and A wave peak velocities). We calculated E/e’ as the ratio of peak early mitral inflow velocity and peak early diastolic mitral annular tissue velocity. As recommended by the American Society of Echocardiography,^22^ we estimated left ventricular mass using a formula and measured diastolic left ventricular diameter and posterior and anteroseptal wall thickness using a leading-edge technique. We calculated the left ventricular h/R ratio as the ratio of the wall thickness (i.e., the sum of left ventricular posterior wall and the left ventricular septum) divided by chamber diameter, and we indexed left ventricular mass to body surface area (using the Du Bois formula).

### Assessment of AVPD

We assessed AVPD using tissue Doppler echocardiography. We measured the displacement of the base of the heart by using the integral of the tissue Doppler s’ wave. The s’ wave, a measure of left ventricular contractility, represents myocardial longitudinal velocity during systole.^23^ Integration of this velocity waveform yields the total AVPD, which corresponds to the movement of the atrioventricular plane toward the apex of the heart during systole. Since the apex of the heart and aortic arch remain stationary during the cardiac cycle,^5–8^ AVPD will be associated with longitudinal stretch of the ascending aorta.^9,24^ Thus, this technique provides an estimate of the change in length of the ascending aorta during the cardiac cycle.

### Aortic stiffness

We signal-averaged and synchronized tonometry waveforms using the electrocardiographic R-wave. We calibrated the signal-averaged brachial pressure waveform peak and trough to the cuff systolic and diastolic pressures, respectively. We calculated mean arterial pressure as the integral of the calibrated brachial pressure waveform.^20^ All other tonometry waveforms were calibrated by using brachial mean and diastolic pressures. We calculated carotid-femoral pulse wave velocity (CFPWV) from tonometry waveforms and body surface measurements that adjusted for parallel transmission in the aortic arch and brachiocephalic artery.^25^

### Clinical evaluation and covariates

Medical history was assessed routinely at each research examination. Age, sex, smoking status, presence of CVD, and use of antihypertensive, hyperlipidemia, and diabetes medications were assessed through questionnaires. Height (meters) and mass (kilograms) were assessed during the examination. Body mass index was calculated as mass in kilograms divided by height in meters squared. Mean arterial blood pressure and heart rate were assessed during tonometry. Serum blood glucose, triglycerides, and cholesterol levels were measured from fasting blood samples.

### Statistical analyses

We tabulated the characteristics of the sample. We inverted CFPWV to limit heteroscedasticity; then we multiplied it by −1000 to convert units to ms/m and rectify the directionality of associations with aortic stiffness. The natural logarithm was used to transform triglycerides, total to high-density lipoprotein cholesterol ratio, fasting blood glucose, body mass index, and E/e’ variables to normalize skewed distributions. Continuous measures were standardized (mean=0, standard deviation=1) for modeling in the primary analysis.

Cross-sectional and longitudinal relations of AVPD, e’, and E/e’ with various CVD risk factors were assessed by using multivariable linear regression. Backward selection was used to identify a parsimonious set of CVD risk factors from among a panel that were separately related to AVPD, e’, and E/e’. We selected CVD risk factors (as covariates and potential confounders) *a priori* based on literature review of reported associations with measures of aortic stiffness or left ventricular diastolic dysfunction and included height, body mass index, total to high-density lipoprotein cholesterol ratio, triglycerides, fasting glucose, heart rate, mean arterial pressure, hypertension treatment, hyperlipidemia treatment, diabetes treatment, current smoking status, and prevalent CVD. Cross-sectional models initially adjusted for age, age^2^, sex, and cohort.

CVD risk factors were added to the model as a group for backward selection. In longitudinal models between examinations, linear regression analysis with backward elimination was used to assess associations of change in AVPD, e’, and E/e’ with changes in a parsimonious set of CVD risk factors. Longitudinal models were adjusted for age, age^2^, sex, cohort, and corresponding baseline AVPD or left ventricular diastolic function measure and baseline CVD risk factor values. The threshold for inclusion and removal from the models was two-sided *P*<0.05.

The foregoing final CVD risk factor models were used to assess cross-sectional and longitudinal relations of AVPD with measures of diastolic function (e’ and E/e’). Expanded cross-sectional models were further adjusted for s’ (expanded model 1) and then left ventricular mass (indexed to body surface area) and left ventricular h/R ratio at visit 1 (expanded model 2). For longitudinal models of change in e’ and E/e’, we adjusted for a base model with CVD risk factors, followed by sequential addition of changes in s’ and AVPD and their corresponding baseline values at visit 1. For both cross-sectional and longitudinal models, we assessed potential effect modification (interaction) by incorporating corresponding interaction terms in the regression models. These included terms for age (above/below median), sex, presence of obesity (body mass index <30 vs. ≥30 kg/m^2^), hypertension treatment, and extent of aortic stiffness (above/below median CFPWV) at visit 1. For significant interactions, we performed stratified analyses. All statistical analyses were performed with SAS version 9.4 for Windows (SAS Institute, Cary, NC), and we considered two-sided *P*<0.05 as statistically significant.

## Results

A flow chart for exclusion of participants is presented in the **Supplemental Figure** and clinical characteristics of included participants at visits 1 and 2 are presented in **Table 1**. The sample at visit 1 comprised relatively healthy middle-aged and older adults, with a preponderance of Generation 3 participants, particularly at visit 2. At visit 2 as compared to visit 1, prevalences of hypertension, hyperlipidemia, and diabetes treatment were higher whereas smoking was less frequent. AVPD, CFPWV, and E/e’ were higher, whereas e’ and s’ were lower at visit 2.

**Table 1.** Clinical characteristics at baseline and follow-up.

| Variable | Visit 1 (N=7117) | Visit 2 (N=4309) |
| --- | --- | --- |
| Age, years | 50±15 | 60±13 |
| Women, n (%) | 3893 (55) | 2393 (56) |
| Offspring participants, n (%) | 2466 (35) | 928 (22) |
| New offspring spouse participants, n (%) | 85 (1) | 45 (1) |
| Omni-1 participants, n (%) | 256 (4) | 145 (3) |
| Generation 3 participants, n (%) | 3921 (55) | 2922 (68) |
| Omni-2 participants, n (%) | 389 (5) | 269 (6) |
| Height, m | 1.7±0.1 | 1.7±0.1 |
| Weight, kg | 78±18 | 81±19 |
| Body mass index, kg/m<sup>2</sup>, median (25th, 75th percentile) | 26.5 (23.4, 30.1) | 27.5 (24.4, 31.5) |
| Total to HDL cholesterol ratio, median (25th, 75th percentile) | 3.4 (2.8, 4.3) | 3.1 (2.5, 3.9) |
| Triglycerides, mg/dL, median (25th, 75th percentile) | 96 (68, 138) | 92 (68, 132) |
| Fasting glucose, mg/dL, median (25th, 75th percentile) | 96 (89, 103) | 97 (91, 105) |
| Heart rate, beats per minute | 61.4±9.7 | 59.5±9.5 |
| Mean arterial pressure, mm Hg | 93±12 | 94±12 |
| Hypertension treatment, n (%) | 1777 (25) | 1378 (32) |
| Hyperlipidemia treatment, n (%) | 1517 (21) | 1326 (31) |
| Diabetes treatment, n (%) | 354 (5) | 312 (7) |
| Currently smoking, n (%) | 854 (12) | 237 (6) |
| Prevalent cardiovascular disease, n (%) | 257 (4) | 170 (4) |
| Aortic spring function and aortic stiffness measures |  |  |
| Atrioventricular plane displacement, cm | 1.53±0.27 | 1.59±0.29 |
| Carotid-femoral pulse wave velocity, m/s<sup>*</sup> | 7.5 (6.5, 9.2) | 8.0 (7.0, 9.6) |
| LV function measures |  |  |
| e', cm/s | 11.2±3.0 | 10.4±2.7 |
| E/e', unitless, median (25th, 75th percentile) | 6.01 (5.09, 7.33) | 6.66 (5.52, 8.20) |
| s', cm/s | 9.6±1.8 | 9.2±1.8 |
| LV h/R ratio, unitless<sup>†</sup> | 0.38±0.06 | --- |
| LV mass index, g/m<sup>†</sup> | 85±19 | --- |

Values are mean±standard deviation, median (25th, 75th percentile), or number (%). HDL, high-density lipoprotein. LV, left ventricular. e', mitral annular early diastolic velocity. E/e', ratio of early mitral inflow velocity and mitral annular early diastolic velocity. s', systolic tissue velocity of the mitral annulus. LV h/R ratio, ratio of the wall thickness (i.e., the sum of LV posterior wall and the LV septum) divided by chamber diameter. LV mass was indexed to body surface area. <sup>\*</sup>N=6877 at visit 1, and N=4253 at visit 2. <sup>†</sup>N=6813 at visit 1; measures were not available at visit 2.

Cross-sectional associations of AVPD, s’, e’, and E/e’ with CVD risk factors at visit 1 are presented in **Table S1**. In models adjusted for age, age^2^, sex, and cohort, e’ tended to be negatively associated and E/e’ tended to be positively associated with CVD risk factors, consistent with worse diastolic function in the presence of higher CVD risk factor burden. AVPD and s’ were generally lower with higher CVD risk factor burden although higher BMI and height were associated with higher AVPD while higher heart rate and height were associated with higher s’. Longitudinal associations of change in (Δ) measures of AVPD and diastolic function with CVD risk factors are presented in **Tables S2 – S4**. In general, an increase in risk factor levels was associated with a reduction in AVPD and e’ and an increase in E/e’. Interestingly, similar to cross-sectional relations (**Table S1**), we observed opposing associations for change in body mass index, which was positively related to change in AVPD while also negatively related to change in e’ and positively related to change in E/e’. The longitudinal CVD risk factor models explained 29%, 64%, and 51% of the variability in AVPD, Δe’, and ΔE/e’, respectively.

Cross-sectional associations of aortic spring measures with left ventricular diastolic function are presented in **Table 2**. Higher AVPD was associated with higher e’ (*β* per SD±standard error=0.43±0.01; *P*<0.001) and lower E/e’ (*β*=-0.35±0.01; *P*<0.001). These associations persisted after further adjustment for differences in left ventricular contractility and structure. Interactions for cross-sectional relations of aortic spring measures with left ventricular diastolic function are summarized in **Table S5**. The association of higher AVPD with higher e’ was stronger among younger participants, women, participants without obesity, participants without prevalent hypertension treatment, and participants with lower aortic stiffness (**Figure 1**). Additionally, the association of higher AVPD with lower E/e’ was stronger among older participants, women, participants with prevalent hypertension treatment, and participants with higher aortic stiffness (**Figure 2**).

**Figure 1.**
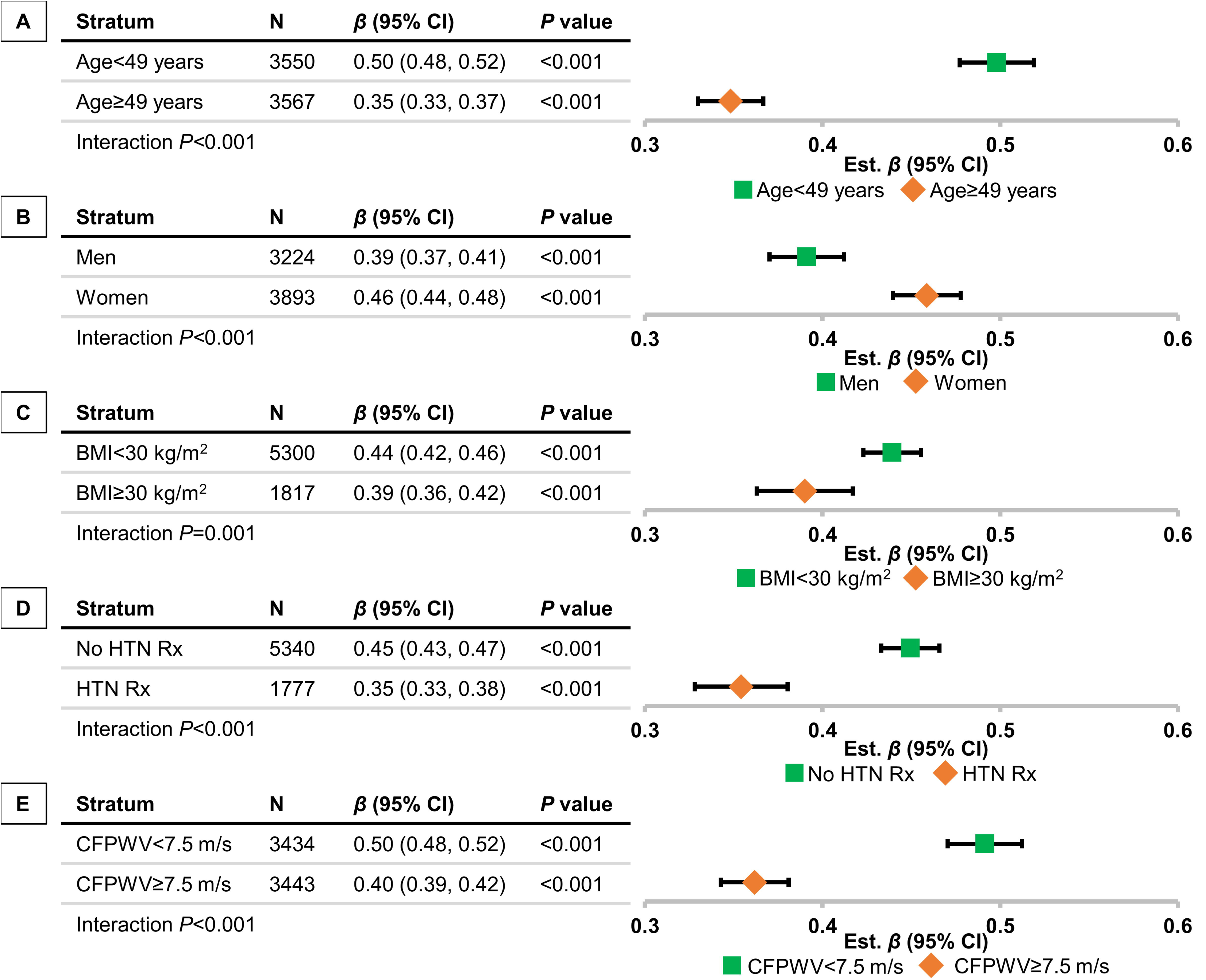
Effect modification for cross-sectional association of atrioventricular plane displacement (AVPD) with e’ by (A) median age, (B) sex, (C) obesity status, (D) presence of hypertension treatment, and (E) extent of aortic stiffness. Effect sizes (*β*s) and 95% CIs from stratified linear regression models. BMI, body mass index. HTN Rx, hypertension treatment. CFPWV, carotid-femoral pulse wave velocity. Age and CFPWV groups were defined as below vs. at/above median at visit 1. Models adjust for age, age^2^, sex (except for sex-stratified models), cohort, height, heart rate, mean arterial pressure, body mass index, total to high-density lipoprotein cholesterol ratio, triglycerides, prevalent hyperlipidemia treatment, and prevalent cardiovascular disease.

**Figure 2.**
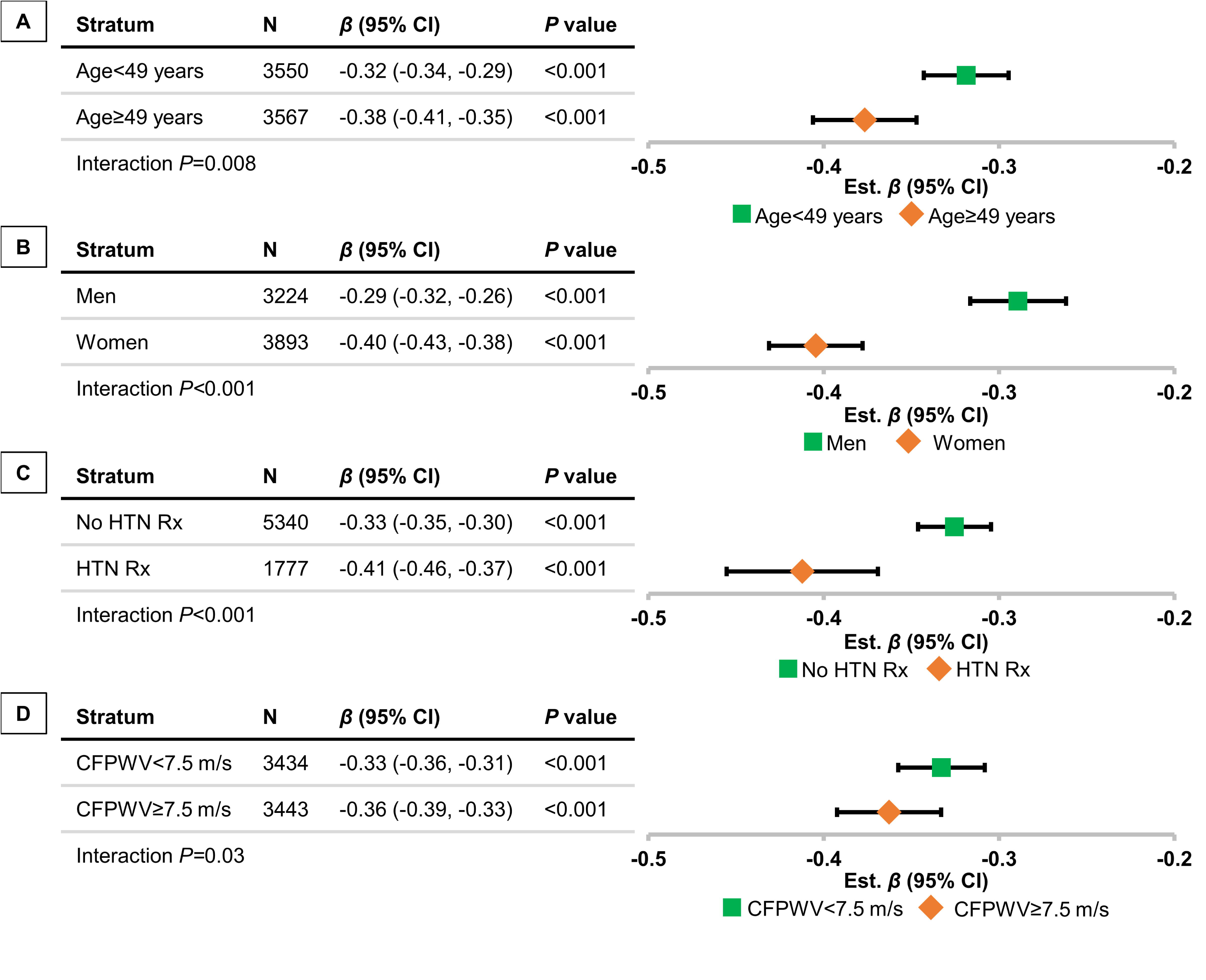
Effect modification for cross-sectional association of atrioventricular plane displacement (AVPD) with E/e’ by (A) median age, (B) sex, (C) presence of hypertension treatment, and (D) extent of aortic stiffness. Effect sizes (*β*s) and 95% CIs from stratified linear regression models. Even when the separate-groups CIs overlap, the CI for differences (interaction) can exclude 0. HTN Rx, hypertension treatment. CFPWV, carotid-femoral pulse wave velocity. Age and CFPWV groups were defined as below vs. at/above median at visit 1. Models adjust for age, age^2^, sex (except for sex-stratified models), cohort, height, heart rate, mean arterial pressure, body mass index, total to high-density lipoprotein cholesterol ratio, prevalent hyperlipidemia treatment, prevalent cardiovascular disease, prevalent diabetes treatment, and prevalent hypertension treatment (except for hypertension treatment-stratified models).

**Table 2.** Cross-sectional relations of atrioventricular plane displacement and left ventricular diastolic function measures at visit 1.

| AVPD models | e' |  | E/e' |  |
| --- | --- | --- | --- | --- |
| | Est. $\beta \pm SE$ (P) | R<sup>2</sup> | Est. $\beta \pm SE$ (P) | R<sup>2</sup> |
| Base model* | 0.43±0.01 (<0.001) | 0.69 | -0.35±0.01 (<0.001) | 0.42 |
| Expanded model 1<sup>†</sup> | 0.53±0.01 (<0.001) | 0.64 | -0.28±0.01 (<0.001) | 0.36 |
| Expanded model 2<sup>‡</sup> | 0.53±0.01 (<0.001) | 0.63 | -0.27±0.01 (<0.001) | 0.36 |

AVPD, atrioventricular plane displacement. e', mitral annular early diastolic velocity. E/e', ratio of early mitral inflow velocity and mitral annular early diastolic velocity. Regression estimates ( $\beta \pm SE$ ) and P values (in parentheses) are 1 standard deviation difference in aortic stretch per 1 standard deviation difference in diastolic function measures. E/e' was natural log transformed. \*Base models for e' are adjusted for age, age<sup>2</sup>, sex, cohort, height, heart rate, mean arterial pressure, body mass index, total to high-density lipoprotein cholesterol ratio, prevalent CVD, triglycerides, and hyperlipidemia treatment; base models for E/e' are adjusted for age, age<sup>2</sup>, sex, cohort, height, heart rate, mean arterial pressure, body mass index, total to high-density lipoprotein cholesterol ratio, prevalent CVD, diabetes treatment, hyperlipidemia treatment, and hypertension treatment. <sup>†</sup>Expanded model 1 further adjusts for s' (myocardial longitudinal velocity during systole). <sup>‡</sup>Expanded model 2 further adjusts for left ventricular (LV) mass (indexed to body surface area) and LV h/R ratio (the ratio of the wall thickness [i.e., the sum of LV posterior wall and the LV septum] divided by chamber diameter).

Associations of longitudinal change in aortic spring measures with change in left ventricular diastolic function between visits are presented in **Table 3**. An increase in AVPD was associated with an increase in e’ (*β*=0.40±0.01; *P*<0.001) and a reduction in E/e’ (*β*=-0.23±0.01; *P*<0.001). These associations persisted after further adjustment for differences in left ventricular contractility and structure. A summary of interactions for longitudinal relations of aortic spring measures with left ventricular diastolic function is presented in **Table S6**. The association of an increase in AVPD with an increase in e’ was stronger among younger participants, participants without obesity, participants without prevalent hypertension treatment, and participants with lower aortic stiffness (**Figure 3**). Additionally, the association of an increase in AVPD with a reduction in E/e’ was stronger among male participants (*β*=-0.23±.02; *P*<0.001 vs. *β*=-0.17±0.02; *P*<0.001; interaction *P*=0.04) and participants with prevalent hypertension treatment (*β*=-0.32±0.05; *P*<0.001 vs. *β*=-0.18±.02; *P*<0.001; interaction *P*=0.02).

**Figure 3.**
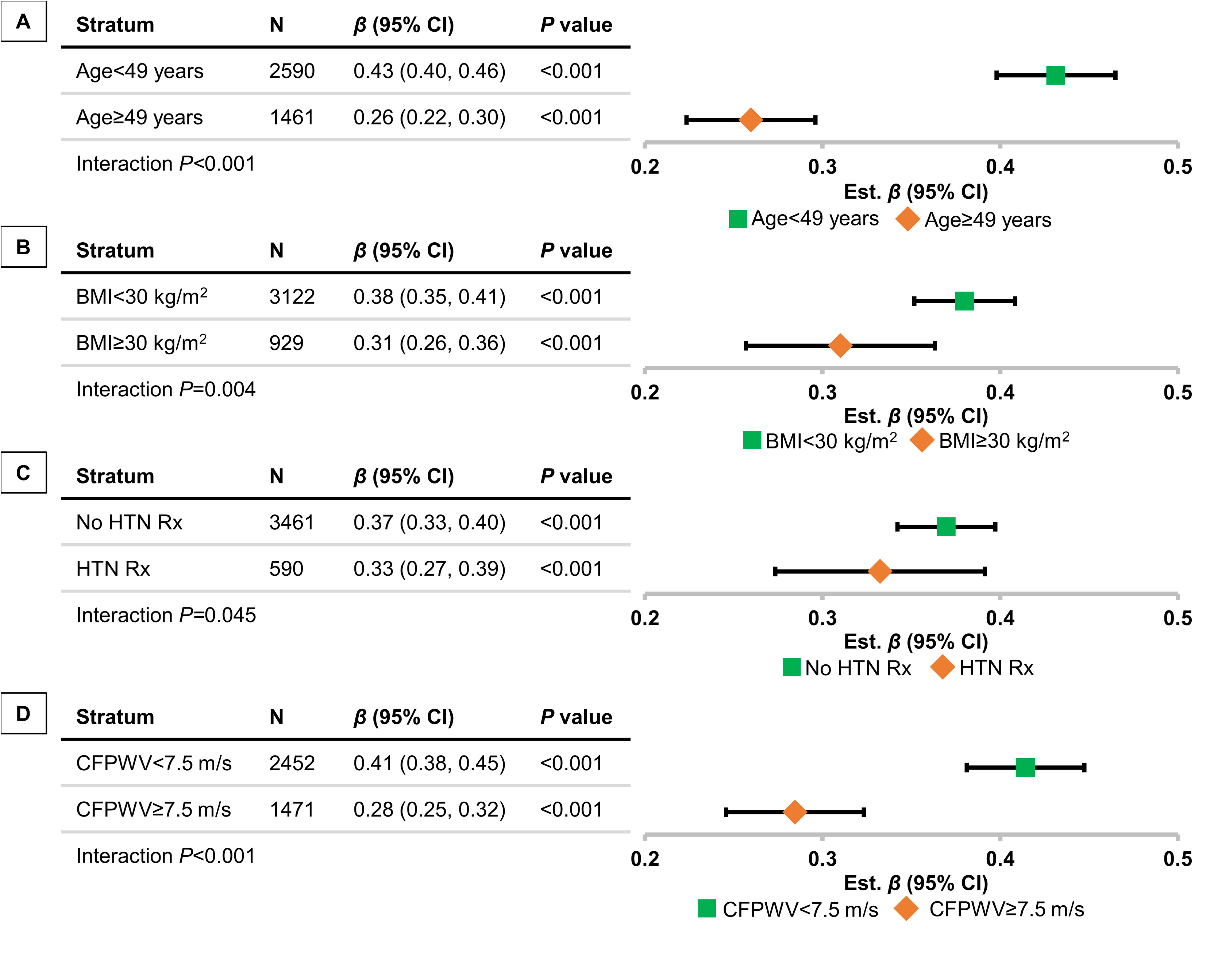
Effect modification for longitudinal associations of ΔAVPD with Δe’ by (A) median age, (B) obesity status, (C) presence of hypertension treatment, and (D) extent of aortic stiffness. AVPD, atrioventricular plane displacement. Effect sizes (*β*s) and 95% CIs from stratified linear regression models. Even when the separate-groups CIs overlap, the CI for differences (interaction) can exclude 0. HTN Rx, hypertension treatment. CFPWV, carotid-femoral pulse wave velocity. Age and CFPWV groups were defined as below vs. at/above median at visit 1. Models adjust for age, age^2^, sex, cohort, height, baseline e’ at visit 1, and baseline and corresponding longitudinal change in cardiovascular disease risk factors (heart rate, mean arterial pressure, body mass index, triglycerides, and smoking status).

**Table 3.** Relations of longitudinal changes in atrioventricular plane displacement with longitudinal changes in left ventricular diastolic function measures (N=4051).

| Model | AVPD variables | $\Delta e'$ | | | $\Delta E/e'$ | | |
| --- | --- | --- | --- | --- | --- | --- | --- |
| | | Est. $\beta \pm SE$ (*P*) | *P* for paired AVPD variables | *R*<sup>2</sup> | Est. $\beta \pm SE$ (*P*) | *P* for paired AVPD variables | *R*<sup>2</sup> |
| Base model* | -- | -- | -- | 0.45 | -- | -- | 0.32 |
| AVPD model<sup>†</sup> | Baseline AVPD | 0.06±0.01 (<0.001) | <0.001 | 0.57 | -0.04±0.02 (0.003) | <0.001 | 0.36 |
| | ΔAVPD | 0.40±0.01 (<0.001) | | | -0.23±0.02 (<0.001) | | |
| Expanded AVPD model 1<sup>‡</sup> | Baseline AVPD | -0.01±0.02 (0.76) | <0.001 | 0.57 | 0.08±0.02 (0.001) | <0.001 | 0.31 |
| | ΔAVPD | 0.40±0.02 (<0.001) | | | -0.15±0.02 (<0.001) | | |
| Expanded AVPD model 2<sup>§</sup> | Baseline AVPD | -0.004±0.02 (0.85) | <0.001 | 0.56 | 0.08±0.02 (0.001) | <0.001 | 0.32 |
| | ΔAVPD | 0.40±0.02 (<0.001) | | | -0.15±0.02 (<0.001) | | |

AVPD, atrioventricular plane displacement. $e'$ , mitral annular early diastolic velocity. $E/e'$ , ratio of early mitral inflow velocity and mitral annular early diastolic velocity. $\Delta s'$ , myocardial longitudinal velocity during systole. $\Delta E/e'$ was natural log transformed. Regression estimates ( $\beta \pm SE$ ) and *P* values (in parentheses) are 1 standard deviation change in aortic stretch per 1 standard deviation change in diastolic function measures. \*Base models include age, age<sup>2</sup>, sex, cohort, height, baseline diastolic function measure (either $e'$ or $E/e'$ at visit 1), and baseline and corresponding longitudinal change in cardiovascular disease risk factors (for $\Delta e'$ : heart rate, mean arterial pressure, body mass index, triglycerides, and smoking status; and for $\Delta E/e'$ : heart rate, mean arterial pressure, body mass index, fasting glucose, and smoking status). <sup>†</sup>AVPD model includes all covariates in the base models as well as ΔAVPD with its corresponding baseline values at visit 1. <sup>‡</sup>Expanded model 1 includes all covariates in the base model as well as ΔAVPD and $\Delta s'$ with their corresponding baseline values at visit 1. <sup>§</sup>Expanded model 2 further adjusts for baseline left ventricular (LV) mass (indexed to body surface area) and LV h/R ratio (the ratio of the wall thickness [i.e., the sum of LV posterior wall and the LV septum] divided by chamber diameter).

## Discussion

### Principal findings

We investigated the cross-sectional and longitudinal relations of AVPD, a surrogate measure of stretch of the ascending aorta and aortic spring function, with measures of left ventricular diastolic function across a broad age range in participants in the community-based FHS. In cross-sectional analyses, higher AVPD was associated with better left ventricular diastolic function (higher e’ and lower E/e’) in models that adjusted for CVD risk factors. Relations persisted in models that further adjusted for left ventricular contractility (s’ velocity)^23^ as well as measures of left ventricular mass and concentric remodeling. We observed evidence of effect modification by age, sex, obesity status (for e’ only), presence of hypertension treatment, and extent of aortic stiffness for these foregoing associations. In longitudinal analyses, an increase in AVPD between examinations was associated with an increase in e’ and a reduction in E/e’, meaning that a reduction in AVPD between visits was associated with worsening left ventricular diastolic function. The longitudinal association of an increase in AVPD with an increase in e’ was stronger in younger participants, participants without obesity, participants without hypertension treatment, and participants with lower aortic stiffness. Additionally, the longitudinal association of an increase in AVPD with a decrease in E/e’ was stronger among men and participants with hypertension treatment. Thus, our results are consistent with the hypothesis that AVPD, a surrogate for ascending aortic stretch during systole and aortic spring function, plays an important role in maintaining left ventricular diastolic function, with putative effects modified by the extent of aortic stiffness, obesity status, age, and sex.

### Relations of CVD risk factors with aortic spring and diastolic function

The associations of left ventricular diastolic dysfunction with CVD risk factors and CVD events, including HFpEF,^26^ are well established. Consistent with prior reports, we observed significant cross-sectional and longitudinal associations of higher levels or higher change in CVD risk factors with worse left ventricular diastolic function in our community-based sample.

During systole, the heart shortens longitudinally, and the AV plane moves toward a stationary apex.^5–8^ Because the aortic root and proximal ascending aorta are anatomically anchored to the base of the heart, this downward motion stretches the aorta.^6,8^ Consequently, AVPD serves as an indirect indicator of proximal aortic stretch, reflecting the dynamics of the aortic spring mechanism. We observed that higher body mass index was associated with higher AVPD, whereas most other CVD risk factors had opposite relations with this surrogate measure of aortic spring function. Nonetheless, body mass index was associated with lower e’ and higher E/e’ suggesting worse diastolic function of the LV. Despite having higher AVPD, our results suggest that participants with obesity have an attenuated ability to recover energy associated with the inferred higher aortic stretch as enhanced early left ventricular diastolic filling. Higher AVPD may not necessarily be recovered as faster early left ventricular diastolic filling (higher e’), possibly because of adverse effects of obesity on the myocardium.^27–29^ Diastolic relaxation is known to be an active, energy requiring process that can be impaired in the presence of altered calcium kinetics or other abnormalities.^30,31^ Thus, while the aortic spring can store energy to facilitate relaxation and early filling, it cannot obviate the need for intact active relaxation and associated extension of myofibrils in the LV. Alternatively, the aforementioned observation may be attributed to excessive adiposity in the pericardium. In early ventricular diastole, the energy accumulated from the proximal aortic stretch results in aortic recoil, which exerts an upward pull on the base of the heart, resulting in a lengthening force along the left ventricular long axis. Pericardial attachments limit movement at the left ventricular apex and contribute to left ventricular lengthening and early cardiac filling as the aortic spring recoils.^32^ Excessive fat in the pericardium and around the apex of the heart may impose a mechanical limit on the ability of the LV to relax briskly in early diastole even if the stretched ascending aorta is exerting a upward force on the base of the heart.^33,34^ Indeed, we observed that cross-sectional and longitudinal associations of greater AVPD with better early left ventricular relaxation (higher e’ and higher Δe’) were attenuated among participants with obesity.

### Relations of aortic spring function and diastolic function

There is a common belief that left ventricular torsion and recoil (via the elastic protein titin and its associated proteins within the myocardial cells) is mostly responsible for energy storage during systole and that this energy is then automatically released like a recoiling spring to enhance refilling of the ventricle during diastole.^2,35,36^ If left ventricular ejection results in an end-systolic volume that is less than the zero pressure volume of the LV, there will be a modest restorative force that will produce a small amount of suction that will facilitate early filling.^37^ However, the left ventricular pressure-volume relation is relatively flat in the low pressure and volume range, meaning the amount of suction will be quite small.^38,39^ Furthermore, early diastolic filling normally results in a left ventricular volume that is considerably greater than the zero pressure volume.^40,41^ Thus, left ventricular recoil may only account for a small fraction of early diastolic filling. Foundational works showed that an external and antecedent preload is necessary to initiate proper diastolic filling in isolated hearts or elongation of cardiac muscle strips.^42–49^ Thus, the intrinsic cardiac elastic properties alone (i.e., recoil of energy stored internally in titin springs as a result of a preceding systolic contraction) cannot fully account for brisk early diastolic left ventricular filling.^50,51^ In our participants, greater AVPD, which implies greater aortic stretch, was associated with better left ventricular diastolic function. We posit that the aortic spring and left ventricular recoil effects due to titin expansion or untwisting may both contribute to early left ventricular diastolic filling through different yet complementary mechanisms.

The relations of AVPD with e’ were diminished in the presence of a stiffer aorta and among older participants. The force required to stretch the aorta during systole increases with higher aortic stiffness, which stores more energy, but also exerts an increased mechanical load on the LV. A higher load on the LV may limit the recovery of stored aortic wall energy because of adverse effects on myocyte structure and function.^9,32^ Additionally, storing more energy at lower systolic displacement because of a stiffer aortic wall will necessarily result in lower restorative displacement during diastole. Alternatively, aortic elongation in relation to aortic aging and stiffening may reduce the amount of preload on the aorta at the beginning of systole and may thereby limit the amount of actual stretch of and energy stored in the proximal aorta at any given level of AVPD. The cellular environment of the aorta may play a role in the differences we observed in relations of AVPD with e’ by age and extent of aortic stiffness. Animal models of aging show that vascular smooth muscle cells isolated from aged aortas can display a senescent phenotype that contributes to aortic stiffness.^52–54^ Accumulation of hypertrophied or senescent vascular smooth muscle cells in the aorta can markedly increase wall viscosity, which can limit aortic stretch during systole. Additionally, viscous losses will increase the dissipation of some of the stored energy (due to aortic stretch) as heat rather than as recovery of elastic recoil and enhanced left ventricular diastolic function.^55^

Interestingly, we observed different interactions (by median age and extent of aortic stiffness) for the cross-sectional relations of AVPD with e’ compared with E/e’. We posit that the association of higher AVPD and presumably higher aortic stretch with higher e’ is stronger in younger individuals and persons with lower aortic stiffness due to their better intrinsic myocardial relaxation capacity. These observations align with well-characterized physiological mechanisms where prolonged calcium transits and delayed cross-bridge dissociation can blur the lines between late systole and early diastole in older individuals and persons with high aortic stiffness, leading to slowed ventricular relaxation and reduced early filling even in the presence of considerable filling pressures or extension forces from a stretched aorta. It is also important to note that while diastole is an active, energy-dependent process, myocardial tissue cannot generate elongation force to expand the chamber. Rather, it must release tension and permit external forces to drive filling. When relaxation is impaired, the ability of the LV to effectively respond to these external diastolic elongation forces is diminished. Conversely, the association of higher aortic stretch with lower E/e’ is stronger in older individuals and persons with higher aortic stiffness presumably because higher left ventricular filling pressure supports a relatively higher mitral inflow (E) at lower levels of AVPD in these groups.

HFpEF is also associated with aortic stiffness.^56,57^ However, we have shown that nonlinear age relations as well as potential period and cohort effects may introduce bias into cross-sectional predictions of longitudinal changes in vascular function.^58^ Therefore, we extended our investigation to assess longitudinal relations with repeated measurements of aortic spring function, CVD risk factors, and left ventricular diastolic function to explore the inter-relations among these measures, and their relation with left ventricular diastolic function and putative contributions to HFpEF. Our findings suggest that left ventricular diastolic early filling is not enhanced as much by a given level of aortic stretch in the presence of high aortic stiffness. This observation implicates the overlapping roles of abnormal mechanical and hemodynamic load on left ventricular diastolic dysfunction. Additional studies that investigate the complex mechanisms and phenotypic diversity of HFpEF and antecedent diastolic dysfunction are warranted to inform better preventative and therapeutic strategies to reduce the incidence and mortality of heart failure.

Contrary to the cross-sectional models, we observed blunted recovery of aortic stretch (as an effect on ΔE/e’) in women in longitudinal models. Our observations are consistent with cross-sectional observations in the AGES cohort where the association of higher stretch-related aortic work with higher early diastolic filling was only observed in men.^9^ Compared to the current study, the AGES participants were significantly older and comprised a narrower age range.

Additionally, women as compared to men in AGES had much lower early ventricular filling (assessed as early diastolic left ventricular filling volume on cardiac magnetic resonance imaging). Furthermore, women are more susceptible to the development of HFpEF.^59,60^ The prior results from AGES among older participants and the current results from FHS support the hypothesis that women may progressively lose their remarkable ability to recapture energy stored in the aortic spring (as observed at visit 1), which might potentially contribute to their higher propensity for HFpEF. Previous studies have shown that women had higher left ventricular stiffness,^61^ slower left ventricular relaxation,^62^ and a greater decline in diastolic long-axis myocardial velocities with increasing age,^63^ which may limit the ability of stored aortic spring energy to facilitate early diastolic filling. Further studies investigating vascular mechanisms underlying sex differences in HFpEF are needed.

## Limitations

Our study has limitations that should be considered. Although we used a prospective design with cross-sectional as well as longitudinal models, all associations of aortic spring measures with measures of left ventricular diastolic function were observational and cannot establish causality. We cannot dismiss the possibility of residual confounding by unknown or unmeasured risk factors (including differences in biochemical and cellular factors like titin and activity and left ventricular collagen deposition). Across exam visits, AVPD increased slightly while diastolic function worsened (e’ decrease while E/e’ increased). From these data, one may infer that AVPD may not actually represent aortic stretch (and subsequent recoil capacity) but rather the reversal of compression of the ascending aorta, particularly in the setting of advanced aortic lengthening and wall stiffening. Furthermore, this lengthened or stiffened aorta would impede rather than facilitate filling. While these small changes may seem paradoxical, they may not be clinically relevant and compare differences of absolute means rather than means adjusted for longitudinal changes in CVD risk factors. Importantly, the direction of the primary association is consistent in cross-sectional and longitudinal analyses.

We cannot dismiss that ceiling effects could have contributed to observed interactions; however, since the interactions between strata and effects within strata are strong and highly significant, it is unlikely that ceiling effects are the primary explanation. Additionally, we observed significant interaction by hypertension treatment for the associations of AVPD with e’ and E/e’. Given that we did not adjust for specific antihypertensive medications as well as the observational nature of our study, we cannot interpret these observations as a drug effect.

However, our data suggest that more severe hypertension (and corresponding higher hypertension treatment) may affect the relations. We acknowledge that gender norms (and associated health behaviors) may contribute to observed differences in associations by sex. Since we did not directly track the displacement of the sinotubular junction, we cannot distinguish between the effects of elongation of the aortic cusps versus elongation of the ascending aorta stretch. However, the methods used for the present study highlight the utility of ultrasound in assessing aortic spring measures compared to magnetic resonance imaging, which is expensive and not as accessible. Our study is susceptible to type-1 error since we did not adjust for multiple testing. Yet, we observed strong associations in the primary analyses and secondary stratified analyses to assess effect modification that would survive multiple testing adjustments. Our samples were mostly composed of White individuals of European ancestry; therefore, our findings may not be generalizable to other ethnic or racial groups.

## Conclusion

In a sample of FHS participants, we observed that higher AVPD, implying greater aortic spring engagement (greater aortic stretch), was related to better left ventricular diastolic function, and favorable changes in aortic spring function were associated with similarly favorable changes in left ventricular diastolic function. Importantly, relations of AVPD with early diastolic left ventricular filling (e’) and filling pressure (E/e’) were independent of measures of contractility (s’). Our findings support the hypothesis that mechanical coupling between the LV and ascending aorta plays a critical role in optimal early diastolic elongation and filling of the LV. Impairment of aortic spring function due to stiffening or elongation of the aorta or reduced AVPD due to reduced longitudinal shortening of the LV during systole, may contribute to the progression of left ventricular diastolic dysfunction. Moreover, elevated aortic stiffness, obesity, aging, and female sex attenuate this mechanism over time, which may make these groups more susceptible to left ventricular diastolic dysfunction and subsequent heart failure, particularly HFpEF. Further research on the mechanical and hemodynamic mechanisms that precede diastolic dysfunction is warranted to inform better therapeutic and preventative strategies.

## Data Availability

The procedure for requesting data from the FHS can be found at https://www.framinghamheartstudy.org/.

## Acknowledgements

From the Framingham Heart Study of the National Heart Lung and Blood Institute of the National Institutes of Health and Boston University Chobanian and Avedisian School of Medicine.

## Sources of funding

This work was supported by the National Heart, Lung, and Blood Institute through contracts [N01-HC-25195, HHSN268201500001I, 75N92019D00031 (Vasan)] and grants [HL071039, HL077447, HL080124, HL093328, HL104184, HL107385, HL126136, HL142983, AG079390, HL131532, HL143227 (Vasan, Mitchell); HL04334 (Vasan); HL073551, HL094898 (Mitchell); AG028321, HL060040, AG066010, HL070100, HL076784, HL092577, HL128914 (Benjamin); 2U54HL120163, U54HL120163 (Hamburg, Benjamin); HL115391, HL168889 (Hamburg); K01HL161494 (Cooper)]. The American Heart Association supported this work through grants 18SFRN34110082 (Benjamin) and 20SRFRN35120118 (Hamburg). The National Institute of Diabetes and Digestive and Kidney Diseases supported this work through grants DK082447 (Mitchell) and DK080739 (Vasan). Dr. Vasan was supported in part by the Evans Medical Foundation and the Jay and Louis Coffman Endowment from the Department of Medicine, Boston University Chobanian & Avedisian School of Medicine.

## Disclosures

Dr. Mitchell is owner of Cardiovascular Engineering, Inc., a company that designs and manufactures devices that measure vascular stiffness. The company uses these devices in clinical trials that evaluate the effects of diseases and interventions on vascular stiffness. G.F.M. also serves as a consultant to and receives grants and honoraria from Novartis, Merck, Bayer, Servier, Philips, and deCODE genetics. The remaining authors have no disclosures to report.

## Notes

### Author Declarations

All protocols were approved by Boston University Medical Center's Institutional Review Board (IRB), and all participants provided written informed consent. The IRB of Boston University Medical Center gave ethical approval for this work.

