## SUPPLEMENTAL APPENDIX for "Interrelations of aortic spring function, cardiovascular disease risk factors, and left ventricular diastolic function: The Framingham Heart Study"

[1]Biology Department, Vassar College, Poughkeepsie, NY; [2]Boston University and NHLBI's Framingham Study, Framingham, MA; [3]Section of Preventive Medicine and Epidemiology, Department of Medicine, Boston University Chobanian and Avedisian School of Medicine, MA; [4]Department of Biostatistics, Boston University School of Public Health, Boston, MA; [5]Section of Cardiovascular Medicine, Department of Medicine, Boston University Chobanian & Avedisian School of Medicine, Boston, MA; [6]Department of Epidemiology, Boston University School of Public Health; [7]Evans Department of Medicine, [8]Whitaker Cardiovascular Institute, Boston University Chobanian & Avedisian School of Medicine, Boston, MA; and [9]The University of Texas School of Public Health San Antonio, Tx; and [10]The University of Texas Health Science Center, San Antonio, TX; [11]Cardiovascular Engineering, Inc., Needham, MA

**Supplemental Content:**

**Supplemental Figure.** Flow diagram of the selection of the study samples. Several excluded participants engaged in offsite visits with limited examination, especially during the pandemic caused by coronavirus disease 2019 (COVID) at visit 2.

**Table S1.** Cross-sectional relations of atrioventricular plane displacement, s', e', and E/e' with cardiovascular disease risk factors at visit 1 (N=7117).

**Table S2.** Relations of change in atrioventricular plane displacement with changes in cardiovascular disease risk factors between two visits (N=4051).

**Table S3.** Relations of change in e' with change in cardiovascular disease vascular risk factors between two visits (N=4051).

**Table S4.** Relations of change in E/e' with change in cardiovascular disease vascular risk factors between two visits (N=4051).

**Table S5.** Summary of interactions for relations of atrioventricular plane displacement with left ventricular diastolic function at visit 1.

**Table S6.** Summary of interactions for longitudinal relations of atrioventricular plane displacement with left ventricular diastolic function.

This supplementary material has been provided by the authors to give the readers additional information about their work.

**Supplemental Figure.** Flow diagram of the selection of the study samples. Several excluded participants engaged in offsite visits with limited examination, especially during the pandemic caused by coronavirus disease 2019 (COVID) at visit 2.

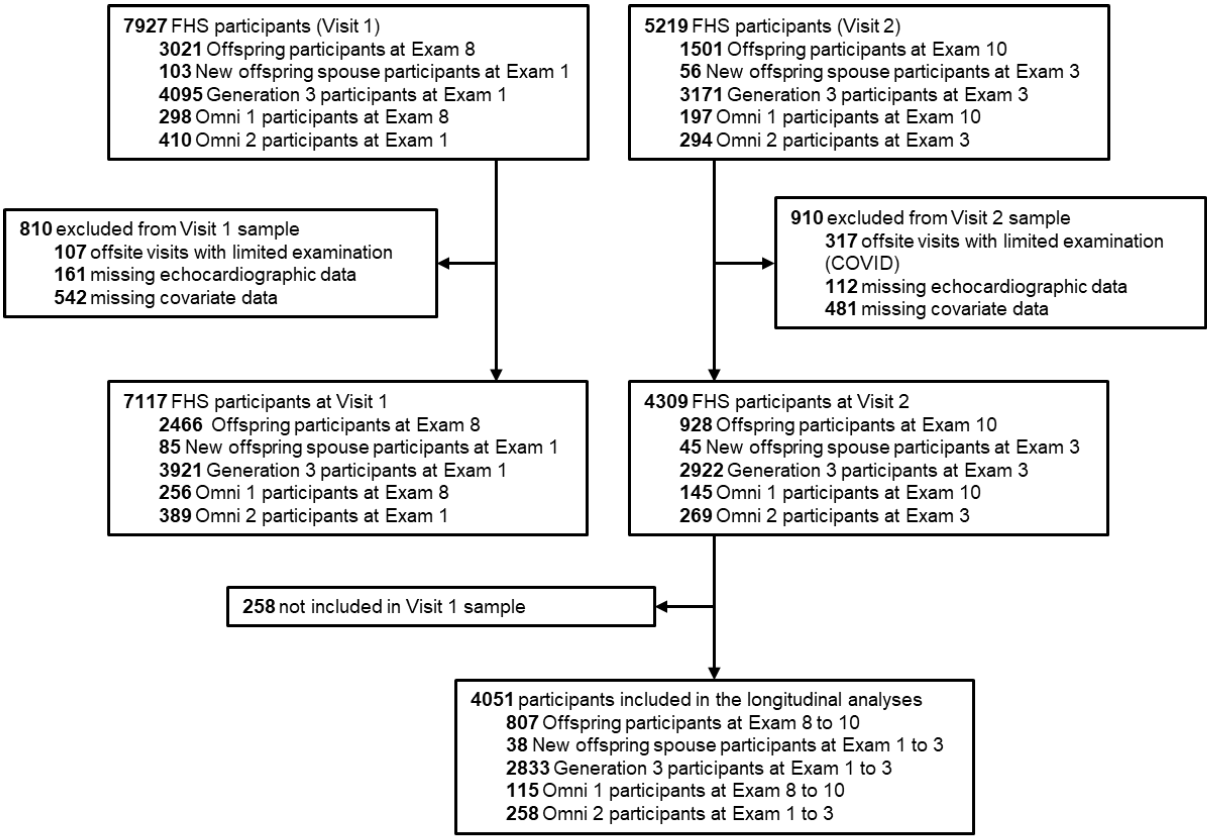

**Table S1.** Cross-sectional relations of atrioventricular plane displacement, s', e', and E/e' with cardiovascular disease risk factors at visit 1 (N=7117).

| | **AVPD** | **s'** | **e'** | **E/e'*** |
|---|---|---|---|---|
| **Variable** | ***β*±SE (*P*)** | ***β*±SE (*P*)** | ***β*±SE (*P*)** | ***β*±SE (*P*)** |
| Height | 0.13±0.02 (<0.001) | 0.18±0.02 (<0.001) | 0.03±0.01 (0.02) | -0.11±0.01 (<0.001) |
| Heart rate | -0.31±0.01 (<0.001) | 0.10±0.02 (<0.001) | -0.14±0.01 (<0.001) | -0.07±0.01 (<0.001) |
| Mean arterial pressure | -0.05±0.01 (<0.001) | -0.13±0.02 (<0.001) | -0.13±0.01 (<0.001) | 0.13±0.01 (<0.001) |
| Body mass index* | 0.14±0.01 (<0.001) | -0.18±0.02 (<0.001) | -0.08±0.01 (<0.001) | 0.18±0.01 (<0.001) |
| Total to HDL cholesterol ratio* | -0.09±0.01 (<0.001) | -0.06±0.02 (<0.001) | -0.05±0.01 (<0.001) | 0.03±0.01 (0.02) |
| Fasting glucose* | -0.03±0.01 (0.01) | -- | -- | -- |
| Prevalent CVD | -0.28±0.06 (<0.001) | -0.38±0.16 (0.02) | -0.14±0.05 (0.003) | 0.19±0.06 (0.001) |
| Current smoking | -- | -0.12±0.04 (0.004) | -- | -- |
| Diabetes treatment | -- | -0.25±0.10 (0.02) | -- | 0.28±0.05 (<0.001) |
| Triglycerides* | -- | -- | -0.03±0.01 (0.01) | -- |
| Hyperlipidemia treatment | -- | -- | -0.07±0.02 (0.005) | 0.08±0.03 (0.01) |
| Hypertension treatment | -- | -- | -- | 0.09±0.03 (0.001) |

AVPD, atrioventricular plane displacement. CVD, cardiovascular disease. HDL, high-density lipoprotein. Regression estimates ($\beta$±SE) and *P* values (in parentheses) for CVD risk factors represent regression slope per 1 standard deviation difference for continuous variables or presence of categorical variables. *Natural log transformed. All models were additionally adjusted for age, age$^2$, sex, and cohort. The threshold to remain in the models was $P<0.05$.

**Table S2.** Relations of change in atrioventricular plane displacement with changes in cardiovascular disease risk factors between two visits (N=4051).

| Variable | Est. $\beta$±SE (*P*) for baseline CVD risk factor variable | Est. $\beta$±SE (*P*) for change in CVD risk factor variable | *P* for paired CVD risk factors |
|---|---|---|---|
| Baseline AVPD | 0.35±0.01 (<0.001) | --- | --- |
| Omni-1 cohort | -0.04±0.09 (0.62) | --- | --- |
| Generation 3 cohort | 0.25±0.05 (<0.001) | --- | --- |
| Omni-2 cohort | 0.19±0.07 (0.01) | --- | --- |
| New offspring cohort | 0.25±0.14 (0.08) | --- | --- |
| Female sex | -0.03±0.04 (0.46) | --- | --- |
| Baseline age | -0.13±0.02 (<0.001) | --- | --- |
| Baseline age$^2$ | 0.01±0.02 (0.45) | --- | --- |
| Baseline height | 0.06±0.02 (0.003) | --- | --- |
| Heart rate | -0.20±0.02 (<0.001) | -0.26±0.02 (<0.001) | <0.001 |
| Body mass index* | 0.06±0.02 (<0.001) | 0.04±0.02 (0.005) | <0.001 |
| Total to HDL cholesterol ratio* | 0.06±0.02 (0.003) | 0.03±0.02 (0.04) | 0.006 |
| Triglycerides* | 0.02±0.02 (0.26) | -0.05±0.02 (0.01) | <0.001 |
| Smoking status | -0.17±0.06 (0.004) | -0.10±0.07 (0.16) | 0.01 |
| **Backward elimination selection†** | | | |
| Base model $R^2$ | 0.22 | | |
| Full model $R^2$ | 0.29 | | |
| Final model $R^2$ | 0.29 | | |

CVD, cardiovascular disease. AVPD, atrioventricular plane displacement. HDL, high-density lipoprotein. *Natural log transformed. Regression estimates ($\beta$±SE) and *P* values (in parentheses) for CVD risk factors are per 1 standard deviation difference in continuous variables or presence of categorical variables. †Base models were adjusted for age, age$^2$, sex, cohort (Offspring cohort as the reference), and baseline AVPD. All risk factor candidates were added and considered for elimination as baseline and ΔCVD risk factor pairs (except height); the threshold to remain in the models was $P<0.05$ for paired CVD risk factors. Variables that were eliminated from the model included weight, fasting glucose, diabetes treatment, hypertension treatment, hyperlipidemia treatment, prevalent CVD, and mean arterial pressure.

**Table S3.** Relations of change in e' with change in cardiovascular disease vascular risk factors between two visits (N=4051).

| Variable | Est. $\beta$±SE (*P*) for baseline CVD risk factor variable | Est. $\beta$±SE (*P*) for change in CVD risk factor variable | *P* for paired CVD risk factors |
|---|---|---|---|
| Baseline e' | 0.34±0.01 (<0.001) | --- | --- |
| Omni-1 cohort | -0.01±0.06 (0.82) | --- | --- |
| Generation 3 cohort | 0.45±0.04 (<0.001) | --- | --- |
| Omni-2 cohort | 0.44±0.05 (<0.001) | --- | --- |
| New offspring cohort | 0.40±0.10 (<0.001) | --- | --- |
| Female sex | -0.06±0.03 (0.02) | --- | --- |
| Baseline age | -0.30±0.02 (<0.001) | --- | --- |
| Baseline age$^2$ | 0.08±0.01 (<0.001) | --- | --- |
| Baseline height | 0.03±0.01 (0.01) | --- | --- |
| Heart rate | -0.11±0.01 (<0.001) | -0.12±0.01 (<0.001) | <0.001 |
| Mean arterial pressure | -0.12±0.01 (<0.001) | -0.11±0.01 (<0.001) | <0.001 |
| Body mass index | 0.01±0.01 (0.52) | -0.05±0.01 (<0.001) | <0.001 |
| Triglycerides* | -0.04±0.01 (0.003) | -0.07±0.01 (<0.001) | <0.001 |
| Smoking status | -0.13±0.04 (0.002) | -0.05±0.05 (0.28) | 0.003 |
| **Backward elimination selection†** | | | |
| Base model $R^2$ | 0.59 | | |
| Full model $R^2$ | 0.64 | | |
| Final model $R^2$ | 0.64 | | |

CVD, cardiovascular disease. *Natural log transformed. Regression estimates ($\beta$±SE) and *P* values (in parentheses) for CVD risk factors are per 1 standard deviation difference in continuous variables or presence of categorical variables. †Base models were adjusted for age, age$^2$, sex, cohort (Offspring cohort as the reference), and baseline e'. All risk factor candidates were added and considered for elimination as baseline and ΔCVD risk factor pairs (except height); the threshold to remain in the models was $P$<0.05 for paired CVD risk factors. Variables that were eliminated from the model included weight, fasting glucose, total to high-density lipoprotein cholesterol ratio, diabetes treatment, hypertension treatment, hyperlipidemia treatment, and prevalent CVD.

**Table S4.** Relations of change in E/e' with change in cardiovascular disease vascular risk factors between two visits (N=4051).

| Variable | Est. $\beta$±SE (*P*) for baseline CVD risk factor variable | Est. $\beta$±SE (*P*) for change in CVD risk factor variable | *P* for paired CVD risk factors |
|---|---|---|---|
| Baseline E/e'* | 0.41±0.01 (<0.001) | --- | --- |
| Omni-1 cohort | -0.09±0.07 (0.21) | --- | --- |
| Generation 3 cohort | -0.45±0.04 (<0.001) | --- | --- |
| Omni-2 cohort | -0.47±0.06 (<0.001) | --- | --- |
| New offspring cohort | -0.29±0.12 (0.01) | --- | --- |
| Female sex | 0.18±0.03 (<0.001) | --- | --- |
| Baseline age | 0.26±0.02 (<0.001) | --- | --- |
| Baseline age$^2$ | 0.08±0.02 (<0.001) | --- | --- |
| Baseline height | -0.07±0.02 (<0.001) | --- | --- |
| Heart rate | 0.02±0.01 (0.24) | -0.06±0.01 (<0.001) | <0.001 |
| Mean arterial pressure | 0.11±0.02 (<0.001) | 0.11±0.01 (<0.001) | <0.001 |
| Body mass index* | -0.00±0.01 (0.87) | 0.10±0.01 (<0.001) | <0.001 |
| Fasting glucose* | 0.04±0.01 (0.007) | -0.00±0.01 (0.73) | 0.01 |
| Smoking status | 0.10±0.05 (0.05) | -0.01±0.06 (0.85) | 0.01 |
| **Backward elimination selection†** | | | |
| Base model $R^2$ | 0.48 | | |
| Full model $R^2$ | 0.51 | | |
| Final model $R^2$ | 0.51 | | |

CVD, cardiovascular disease. *Natural log transformed. Regression estimates ($\beta$±SE) and *P* values (in parentheses) for CVD risk factors are per 1 standard deviation difference in continuous variables or presence of categorical variables. †Base models were adjusted for age, age$^2$, sex, cohort (Offspring cohort as the reference), and baseline E/e'. All risk factor candidates were added and considered for elimination as baseline and ΔCVD risk factor pairs (except height); the threshold to remain in the models was $P<0.05$ for paired CVD risk factors. Variables that were eliminated from the model included weight, triglycerides, total to high-density lipoprotein cholesterol ratio, diabetes treatment, hypertension treatment, hyperlipidemia treatment, and prevalent CVD.

**Table S5.** Summary of interactions for relations of atrioventricular plane displacement with left ventricular diastolic function at visit 1.

| | e' | E/e' |
|---|---|---|
| **Variable** | *P* Value | *P* Value |
| Interaction by above median age | <0.001 | 0.008 |
| Interaction by sex | <0.001 | <0.001 |
| Interaction by presence of obesity | 0.001 | 0.16 |
| Interaction by above median CFPWV | <0.001 | 0.03 |
| Interaction by hypertension treatment | <0.001 | <0.001 |

e', mitral annular early diastolic velocity. E/e', ratio of early mitral inflow velocity and mitral annular early diastolic velocity. CFPWV, carotid-femoral pulse wave velocity. *P*-values for interaction terms are presented. E/e' was natural log transformed. All e' models are adjusted for age, age$^2$, sex, cohort, height, body mass index, heart rate, mean arterial pressure, total to high-density lipoprotein cholesterol ratio, prevalent CVD, triglycerides, and hyperlipidemia treatment. All E/e' models are adjusted for age, age$^2$, sex, cohort, height, body mass index, heart rate, mean arterial pressure, total to high-density lipoprotein cholesterol ratio, prevalent CVD, diabetes treatment, hyperlipidemia treatment, and hypertension treatment.

**Table S6.** Summary of interactions for longitudinal relations of atrioventricular plane displacement with left ventricular diastolic function.

| | Δe’ | ΔE/e’ |
|---|---|---|
| **Variable** | *P* Value | *P* Value |
| Interaction by above median age | <0.001 | 0.97 |
| Interaction by sex | 0.10 | 0.04 |
| Interaction by presence of obesity | 0.004 | 0.54 |
| Interaction by above median CFPWV | <0.001 | 0.80 |
| Interaction by hypertension treatment | 0.045 | 0.02 |

e’, mitral annular early diastolic velocity. E/e’, ratio of early mitral inflow velocity and mitral annular early diastolic velocity. CFPWV, carotid-femoral pulse wave velocity. *P*-values for interaction terms are presented. ΔE/e’ was natural log transformed. All models include age, age$^2$, sex, cohort, height, baseline diastolic function measure (either e’ or E/e’ at visit 1), and baseline and corresponding longitudinal change in cardiovascular disease risk factors (for Δe’: heart rate, mean arterial pressure, body mass index, triglycerides, and smoking status; and for ΔE/e’: heart rate, mean arterial pressure, body mass index, fasting glucose, and smoking status).
